# At-home wearables and machine learning capture motor impairment and progression in adult ataxias

**DOI:** 10.1101/2024.10.27.24316161

**Authors:** Rohin Manohar, Faye X. Yang, Christopher D. Stephen, Jeremy D. Schmahmann, Nicole M. Eklund, Anoopum S. Gupta

## Abstract

A significant barrier to developing disease-modifying therapies for spinocerebellar ataxias (SCAs) and multiple system atrophy of the cerebellar type (MSA-C) is the scarcity of tools to sensitively measure disease progression in clinical trials. Wearable sensors worn continuously during natural behavior at home have the potential to produce ecologically valid and precise measures of motor function by leveraging frequent and numerous high-resolution samples of behavior.

Here we test whether movement-building block characteristics (i.e., submovements), obtained from the wrist and ankle during natural behavior at home, can sensitively capture disease progression in SCAs and MSA-C, as recently shown in amyotrophic lateral sclerosis (ALS) and ataxia telangiectasia (A-T).

Remotely collected cross-sectional (*n* = 76) and longitudinal data (*n* = 27) were analyzed from individuals with ataxia (SCAs 1, 2, 3, and 6, MSA-C) and controls. Machine learning models were trained to produce composite outcome measures based on submovement properties. Two models were trained on data from individuals with ataxia to estimate ataxia rating scale scores. Two additional models, previously trained entirely on longitudinal ALS data to optimize sensitivity to change, were also evaluated.

All composite outcomes from both wrist and ankle sensor data had moderate to strong correlations with ataxia rating scales and self-reported function, strongly separated ataxia and control populations, and had high within-week reliability. The composite outcomes trained on longitudinal ALS data most strongly captured disease progression over time.

These data demonstrate that outcome measures based on accelerometers worn at home can accurately capture the ataxia phenotype and sensitively measure disease progression. This assessment approach is scalable and can be used in clinical or research settings with relatively low individual burden.

## Introduction

Disease-modifying therapies are under development for neurodegenerative diseases, including spinocerebellar ataxias (SCAs)^1–4^ and multiple system atrophy (MSA).^5–9^ A significant barrier to the development of therapies is the scarcity of tools that can sensitively measure disease progression and possible treatment response in clinical trials that are limited by size and duration.^9-10^ Clinician-scored disease rating scales, such as the Scale for the Assessment and Rating of Ataxia (SARA)^11^ are the current gold standard. Yet, rating scale sensitivity is limited by several factors, such as rater subjectivity, low granularity of the scoring system, and variability in participant energy level at the time the exam is performed.^10,12^ Rating scales are also prone to floor and ceiling effects,^10,13^ leading to difficulties capturing very mild or severe symptoms. Furthermore, it is unclear if the motor tasks evaluated as part of these scales reflect everyday motor function.^14^

Wearable devices are a promising technology to objectively quantify movement changes present in ataxias. Wrist sensors containing triaxial accelerometers have been used in upper-limb task-based paradigms,^15–25^ such as pronation-supination and finger-to-nose, and have shown that measures derived from the sensor data can distinguish between ataxia and control participants and correlate well with ataxia rating scales.^16,25^ Similarly, accelerometers worn on the lower limbs and/or trunk have been used to quantify gait and balance changes during in-clinic or laboratory paradigms, such as the two-minute walk test.^17,19–23,26–29^ Measures including lateral step deviation strongly correlated with SARA scores and captured one-year disease progression more sensitively than SARA.^18,26^

Accelerometers have also been used at home to capture cerebellar gait features in natural environments.^18,30^ Thierfelder *et al.*^30^ used wearable sensors on the feet and lower back to capture turning movements of ataxic participants over a four to six-hour period of unsupervised daily living (including at least 30 minutes of walking) at baseline and after one year.^30^ Lateral velocity change (LVC) and outward acceleration captured dynamic balance control during turning. These features correlated with SARA scores, were able to capture changes in pre-ataxic subjects, and were sensitive to disease progression.^30^

More recently, accelerometers have also been used in ataxia populations to capture behavior continuously at home for multiple days without imposing requirements on the individual’s behavior.^31–35^ The absence of behavioral requirements reduces barriers to frequent or continuous data collection over time, facilitates participation from young children and more severely affected individuals, and has the potential to generate measures that more closely represent daily function.^33–35^

For example, Fichera *et al.*^31^ used accelerometers worn on the wrist and lower back to capture the natural behavior of Friedreich’s ataxia participants for one week at baseline and one year later. Between the two timepoints, they observed a decrease in the vector magnitude of all axes (VM3) and per-minute step count, along with an increase in sedentary periods. Both waist-sensor VM3 and the percentage of time spent sedentary were significantly correlated with SARA gait subscore and captured disease progression over one year.^31^ In another Friedreich’s ataxia study,^32^ participants wore an accelerometer on their dominant tricep for five days at baseline and again at six-month and one-year intervals. Daily step count was more sensitive to disease progression than clinical outcomes (SARA, Friedreich Ataxia Rating Scale Upright Stability Subscale) at six months, but FARS USS had greater effect sizes at one year. The authors noted that large variability in daily step count, influenced by seasonal effects, lifestyle changes, and physical activity enjoyment may limit its efficacy in clinical trials.^32^

Submovement-based analysis is another approach for quantifying ataxia severity based on natural behavior at home.^34^ Submovements are theorized to be building blocks for movements,^36^ and are typically characterized by bell-shaped velocity curves. There is evidence that the decomposition of movement observed in the ataxia phenotype^37^ is reflected at the submovement level for upper and lower limbs, during task-based^25,28,38^ and task-free^33-34,39^ assessment paradigms, with submovements becoming smaller, slower, and less regular with increasing ataxia severity.

We previously showed that submovement-based measures, derived from accelerometer data collected continuously at-home from the ankle and wrist in individuals with SCAs and MSA-C, were highly reliable, correlated strongly with self-reported function and ataxia rating scales, and distinguished between ataxia and control participants.^33^ Here, we evaluate cross-sectional properties in an expanded cohort and report longitudinal follow-up data to test the hypothesis that submovement measures sensitively capture disease progression in adult ataxias during at-home, remote monitoring.

## Materials and methods

### Recruitment and consent

Participants took part in the study between November 2019 and March 2024. The study was approved by the Partners Healthcare Research Committee Institutional Review Board (no. 2019P003458). Age-matched control data collected under a separate study (no. 2019P002752) was also used in this analysis to increase the size of the control cohort. Informed consent was obtained from all subjects before participation in each research study according to the Declaration of Helsinki. Study participants were recruited from the Massachusetts General Hospital (MGH) Ataxia Center, as well as through the National Ataxia Foundation (NAF) and Friedreich’s Ataxia Research Alliance (FARA) websites and newsletters. Participants’ spouses were recruited as controls if they had no known ataxia risk factors, movement disorders, significant orthopedic conditions, or other conditions that could affect motor ability. Relatives of participants were allowed to participate as controls only if they received a negative genetic test result for ataxia. Other controls were either friends of a current participant or were recruited from the NAF or FARA websites. To participate in the study, all subjects had to be: (i) native English speakers, (ii) at least 18 years old, (iii) able to use a computer mouse, (iv) able to walk without another human’s assistance (assistive devices such as canes and walkers were allowed). SCA participants were required to have genetic confirmation of their diagnosis or a first-degree relative with genetically confirmed SCA and a phenotype consistent with this diagnosis without an alternative explanation. Individuals with MSA-C with a probable or possible diagnosis based on the 2008 diagnostic criteria^40^ were eligible to participate. Participants older than 80 were excluded from analysis to reduce the heterogeneity of the cohort and factors related to aging.

76 individuals were included in the analysis: 24 SCA3 (seven pre-ataxic), 10 SCA6 (four pre-ataxic), six SCA1, four SCA2 (two pre-ataxic), seven MSA-C, and 25 controls. Of the 25 controls, 14 were from a different study following similar procedures, referred to as the second control cohort. Participants were labeled pre-ataxic if at any point during the study they had a SARA score < 3.^41^ Of the 13 pre-ataxic participants, at least two individuals were also pre-symptomatic, defined as not having any symptoms clearly attributable to their genetic diagnosis of SCA. One additional individual had infrequent episodes of vertigo, but did not have symptoms outside of the episodes. Pre-symptomatic classification was made based on review of their current neurologist note; however, due to the small number of individuals and subjectivity with classification, this group was included with pre-ataxic individuals and not analyzed as a separate group. Six MSA-C participants had a probable diagnosis of MSA, and one had a possible diagnosis. Participants came from a broad geographic background, living in 25 different states nationwide. Individual-level demographic and clinical information are shown in Supplementary Table 1.

Of the 76 individuals, 35 had longitudinal data across two or more timepoints: 14 SCA3 (five pre-ataxic), four SCA1, four SCA6 (two pre-ataxic), two SCA2 (one pre-ataxic), three MSA-C, and eight controls. For ataxia versus control comparisons, pre-ataxic individuals were analyzed separately for ataxic participants. Analysis was not performed on the pre-ataxic individuals’ longitudinal data due to small sample size. Thus, 27 individuals were included in longitudinal analysis. From November 2019 to January 2023, longitudinal participants were asked to complete the second timepoint 12 months after their initial visit. Beginning in January 2023, all new participants were asked to complete their second timepoint 6 months after the initial visit. Longitudinal participants completed 2.9 visits on average (range 2-4) over an average of 1.7 years (range 0.5-2.8).

Of the 41 study participants without longitudinal data, 17 were not due for follow-up by March 2024 and four participants’ follow-up timepoints were delayed until after March 2024. Of the remaining 20 participants (13 with ataxia, seven controls), three dropped out (two controls were too busy to continue participating, one ataxia participant did not want to continue due to severity), 12 were lost to follow-up, and three passed away (two spouses of the deceased participants also withdrew from the study).

### Equipment and supplies

39 study participants were provided with a study laptop for all of their timepoints, while 37 participants used a personal computer that met the study requirements.^33^ All participants were given two GENEActiv wearable sensors that record triaxial accelerometer data at 100Hz. One device was worn on the dominant ankle, and the other was worn on the dominant wrist. Participants wore both sensors simultaneously for one week as previously described.^33^

### Virtual Appointment and Neurological Assessment

The study coordinator met with participants over Zoom to help turn on and position the GENEActiv devices and conduct a neurological assessment. The 14 participants in the second control cohort did not complete a virtual neurological assessment. The assessment was recorded with the participant’s consent. The study coordinator, who was trained to administer ataxia assessments, or an ataxia-specialist neurologist, administered the Scale for the Assessment and Rating of Ataxia (SARA) and the Brief Ataxia Rating Scale (BARS) half-point version.^11,42,43^ A single ataxia-specialist neurologist (A.S.G.) completed the rating scale scoring from the recordings for every exam. Ratings from a second ataxia-specialist neurologist were not included as the second rater scores were only performed at the first time point.^33^

The sitting component of SARA was excluded due to the inability to perform the task remotely, making the SARA score range 0-36 instead of 0-40. Linear scaling was applied to the total SARA and BARS scores of individuals with missing subscore data (see below) to ensure that the maximum possible score was consistent across all participants.^33^

35 SARA subscores for individual tasks were missing across 15 sessions from 13 participants (nine stance, nine finger-chase right, eight finger-chase left, two heel-to-shin left, two heel-to-shin right, two gait, one alternating hand movements right, one alternating hand movements left, and one finger-nose right). The reasons for the missed scores included poor task performance (20), safety concerns (nine), technical difficulties (four), and environmental constraints (two).

Eight BARS subscores for individual tasks were missing across seven sessions from seven participants (two gait, two heel-to-shin left, two heel-to-shin right, one finger-nose right, and one oculomotor). The reasons for the missed scores included poor task performance (three), technical difficulties (three), and safety concerns (two).

For two participants in the control cohort where remote assessments were performed, no exam was performed for their second timepoint due to scheduling challenges. For one participant, because their timepoint 1 (T1) and timepoint 3 (T3) exam scores were all zeroes, the participant was given all zeros for the timepoint 2 (T2) exam. For the other participant, their total T1 SARA and BARS scores were 3.5 and 1.5, respectively, while their T3 scores were three and one. The only discrepancies were SARA and BARS finger-nose left scores, so the T3 scores were used because T3 occurred closer to T2 than T1 did. This brought the participant’s total T2 SARA and BARS scores to three and one, respectively.

### Questionnaires

SCA study participants completed several Patient-Reported Outcome Measure (PROM) questionnaires at each timepoint (PROM-Ataxia, Dysarthria Impact Scale, Rand 36 Item Short Form Health Survey, five-level EuroQol 5D, and Neurology Quality-of-Life Fatigue Subscale).^44–48^ The only questionnaire used in this analysis was PROM-Ataxia due to interest in comparing wearable measures with ataxia-specific patient-reported measures of function.^44^Question 64 of PROM-Ataxia (“I can think of the words I want to say in conversation”) was excluded from analysis due to incorrect coding in REDCap until April 2023. This question was incorrectly grouped in the prior PROM-Ataxia subsection; thus, the answer choices were presented in reversed order of valence with the wording of “never” - “always” instead of “without any difficulty” - “unable to do.” Although answers were corrected by reversing the scores, this question was removed from the PROM-Ataxia total score for consistency in longitudinal analysis. Thus, all analyses with PROM-Ataxia in this study removed question 64, and a new total score out of the 69 remaining questions was calculated. PROM-Ataxia questions were split into overlapping domain-specific subsections to correlate relevant domains with wrist and ankle sensor data: motor (28), arm (15), and gait and balance (12).^33^

The study initially asked participants to complete all questionnaires twice, separated across four weeks. After February 2023, participants were only asked to complete the questionnaires once at each time point. To ensure consistency across timepoints, for timepoints in which a survey was completed twice, only the first completed survey was used in the analysis.

### Wearable sensor data processing

Participants’ wearable sensor data were manually divided into day and night segments based on changes in each participant’s activity level visualized in the accelerometer data.^33–35^ To account for differences in the time of day that participants started wearing the sensors, day/night segmentation began at sleep onset during the first complete night of recording. This produced a maximum of six consecutive 24-hour periods of recording. Data analysis used daytime segments only.

Continuous wrist and ankle sensor data were processed and analyzed, as previously described^33,34^, to extract submovements (SM) and quantify activity intensity (AI). Six key SM summary features based on the mean and variance of distance, velocity, and acceleration, and one activity intensity feature (entropy of AI) were selected a priori based on their promising cross-sectional properties.^33^ Four machine-learned composite models were applied to the extracted features from each week-long recording session. Each model performed a weighted linear combination of the features to generate an overall estimate of severity for the session. Two of the models were trained and evaluated using cross-validation on this ataxia dataset, one based on wrist sensor data (Ataxia-Wrist) and one on ankle sensor data (Ataxia-Ankle), to predict BARS total score. The other two models were trained on a separate dataset from individuals with ALS, also with one model trained on wrist data (ALS-Wrist) and one on ankle data (ALS-Ankle).^39^ The ALS models were not trained to predict a clinical outcome, but instead were trained on a large longitudinal ALS dataset to predict change over time. We hypothesized that the ALS models, which were optimized to measure progression, may also capture elements of progression in this population of individuals with ataxia.

### Statistical Analysis

Statistical analyses were completed in MATLAB version R2022a (Mathworks, Natick, MA). The Mann-Whitney *U*-test was used to determine sensor measure differences between disease and control groups and Cohen’s *d* was used to measure effect size. The Benjamini-Hochberg method was used to adjust for multiple comparisons and corrected p-values were reported.^49^ Corrected p-values < 0.05 were considered significant. To evaluate reliability of wrist and ankle features, features were computed from data recorded on days 1-3 and days 4-6, separately, and single-measure intraclass correlation coefficients (ICCs) were computed using a two-way mixed effects model.^50^ Pearson correlation coefficients and p-values were used to evaluate the relationship between ankle and wrist sensor features with ataxia rating scales (SARA and BARS) and patient-reported measures of function (PROM-Ataxia). As above, the Benjamini-Hochberg method was used to adjust for multiple comparisons for each sensor type.^49^ To avoid inflated correlation values driven by differences between control and ataxia participants, Pearson correlation coefficients were computed using data from ataxia participants only. For longitudinal data analysis, each participant’s progression rate for a given outcome was determined by fitting a linear regression model to the individual’s longitudinal data for the outcome and using the slope of the curve to represent progression over time.^51^ The mean and standard deviation of the slope for each outcome were computed across the population, and the mean to standard deviation ratio (MSDR) was used to report effect size for sensitivity to change. The Wilcoxon signed-rank test was used to determine if the outcome’s slope was significantly different from zero. Corrected p-values < 0.05 were considered significant.

## Results

### Cross-sectional properties of wrist movement features

All wrist sensor-derived submovement (SM) and activity intensity (AI) features, and composite models were significantly correlated with SARA (and BARS) total score (*|r| =* 0.50 - 0.76, Table 2), SARA arm subscore (*|r|* = 0.35 - 0.57), BARS arm subscore (*|r|* = 0.38 - 0.63), PROM-Ataxia total score (*|r|* = 0.50 - 0.62), and PROM-Ataxia arm subscore (*|r|* = 0.55 - 0.68). The mean and standard deviation of SM distance, velocity, and acceleration all decreased with increasing disease severity. Consistency analysis, comparing movement patterns obtained from the first versus second half of the week, demonstrated high reliability of all features and models (*ICC* = 0.84 - 0.95). The two wrist-trained models, Ataxia-Wrist and ALS-Wrist, demonstrated consistently high correlations with clinical and patient-reported assessments and high reliability (*ICC* = 0.95, Table 2).

**Table I.**
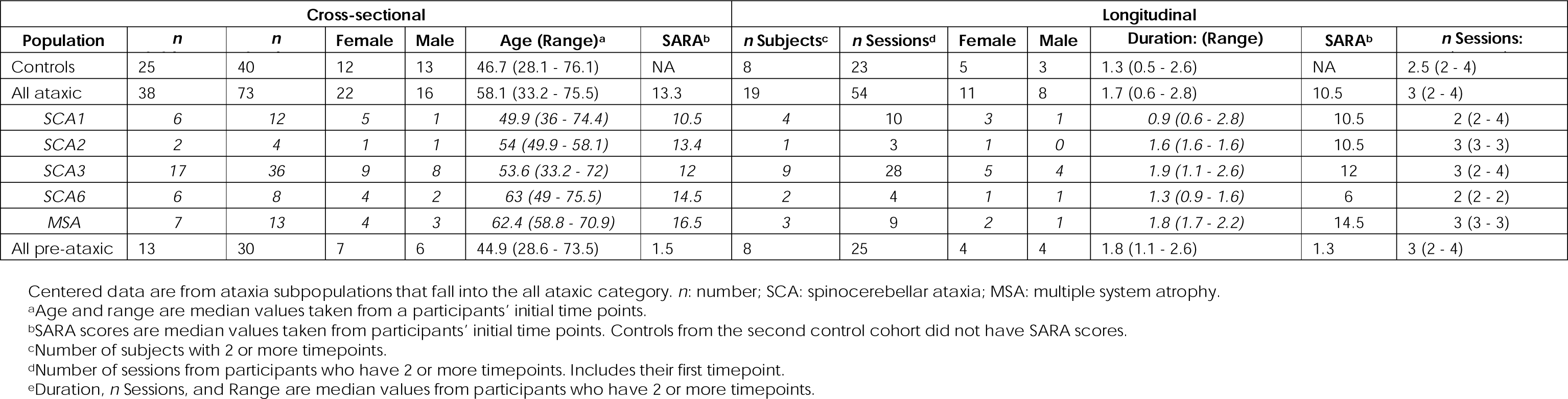
Summary of participant demographic and clinical information.

**Table 2.** Relationship with scales and test-retest reliability of wrist sensor features and models.

| Wrist Feature or Model Name | Statistic | Relationship with SARA (BARS)<sup>a</sup> |  |  |  | Relationship with PROM-Ataxia<sup>a</sup> |  |  |  | Test-retest reliability (N = 138)<sup>b,c</sup> |
| --- | --- | --- | --- | --- | --- | --- | --- | --- | --- | --- |
|  |  | Total (N = 103) |  | Arm subscore (N = |  | Total (N = 103) |  | Arm subscore (N = |  |  |
|  |  | r | p-val | r | p-val | r | p-val | r | p-val | ICC |
| SM Distance<br>*Short SMs PC2* | Mean | -0.62 (-0.59) | 9.2E-12 | -0.45 (-0.49) | 3.5E-06 | -0.57 | 6.2E-10 | -0.67 | 2.2E-14 | 0.90 |
|  | SD | -0.62 (-0.60) | 1.2E-11 | -0.47 (-0.52) | 2.2E-06 | -0.56 | 1.7E-09 | -0.65 | 1.2E-13 | 0.85 |
| SM Velocity<br>*Long SMs PC2* | Mean | -0.50 (-0.48) | 5.5E-08 | -0.35 (-0.38) | 2.8E-04 | -0.52 | 2.1E-08 | -0.63 | 1.8E-12 | 0.92 |
|  | SD | -0.57 (-0.55) | 4.4E-10 | -0.43 (-0.47) | 5.8E-06 | -0.55 | 4.2E-09 | -0.66 | 6.7E-14 | 0.91 |
| SM Acceleration<br>*Long SMs PC2* | Mean | -0.61 (-0.59) | 2.0E-11 | -0.45 (-0.49) | 3.4E-06 | -0.60 | 1.5E-10 | -0.68 | 1.8E-14 | 0.94 |
|  | SD | -0.58 (-0.57) | 1.4E-10 | -0.45 (-0.50) | 3.9E-06 | -0.57 | 6.7E-10 | -0.64 | 3.4E-13 | 0.87 |
| Activity Intensity | Entropy | -0.59 (-0.55) | 6.5E-11 | -0.46 (-0.52) | 2.7E-06 | -0.54 | 8.0E-09 | -0.59 | 4.3E-11 | 0.90 |
| Ataxia Ankle Severity Model |  | 0.55 (0.50) | 1.9E-09 | 0.42 (0.47) | 9.6E-06 | 0.50 | 7.9E-08 | 0.55 | 1.9E-09 | 0.86 |
| Ataxia Wrist Severity Model<sup>d</sup> |  | 0.76 (0.73) | 2.2E-19 | 0.55 (0.63) | 9.7E-09 | 0.62 | 2.7E-11 | 0.68 | 1.9E-14 | 0.95 |
| ALS Ankle Severity Model |  | -0.59 (-0.59) | 6.0E-11 | -0.44 (-0.52) | 3.6E-06 | -0.52 | 1.8E-08 | -0.61 | 9.5E-12 | 0.84 |
| ALS Wrist Severity Model<sup>d</sup> |  | -0.72 (-0.71) | 3.4E-17 | -0.57 (-0.63) | 2.7E-09 | -0.59 | 1.6E-10 | -0.68 | 3.0E-14 | 0.95 |

BARS values represented in parentheses in the “Relationship with SARA” r-values section of the table.
<sup>a</sup>Relationship with clinical scales and PROMs exclude control subjects, whereas test-retest reliability includes controls.
<sup>b</sup>N represents number of sessions for ataxic, pre-ataxic, and control participants.
<sup>c</sup>Test-retest reliability analysis utilizes sessions with at least six full days of data.
<sup>d</sup>Ataxia Wrist and ALS Wrist models are bolded to highlight the strength of their correlations and test-retest reliability compared to other features and models.
SM: Submovement; SD: Standard deviation; PC2: Principal component 2 analysis direction; SARA: Scale for the assessment and rating of ataxia; BARS: Brief Ataxia Rating Scale;
PROM-Ataxia: Patient Reported Outcome Measure of Ataxia; ICC: Intraclass correlation coefficient; r: Pearson correlation coefficient; ALS: amyotrophic lateral sclerosis.

Next, wrist movement features and models were compared between ataxia (excluding pre-ataxic individuals) and control populations. Significant differences were observed between ataxic individuals and controls across all features and models (*e.s.* = 0.6 - 1.7, Table 3). Significant differences were also seen between specific populations (SCA3, SCA1, SCA6, and MSA-C) and the control group. No significant differences were observed between pre-ataxic individuals and control participants. The Ataxia-Wrist and ALS-Wrist models demonstrated consistently strong separation between all ataxic populations and the control group (*e.s.* = 1.3 - 2.7, Table 3).

**Table 3.**
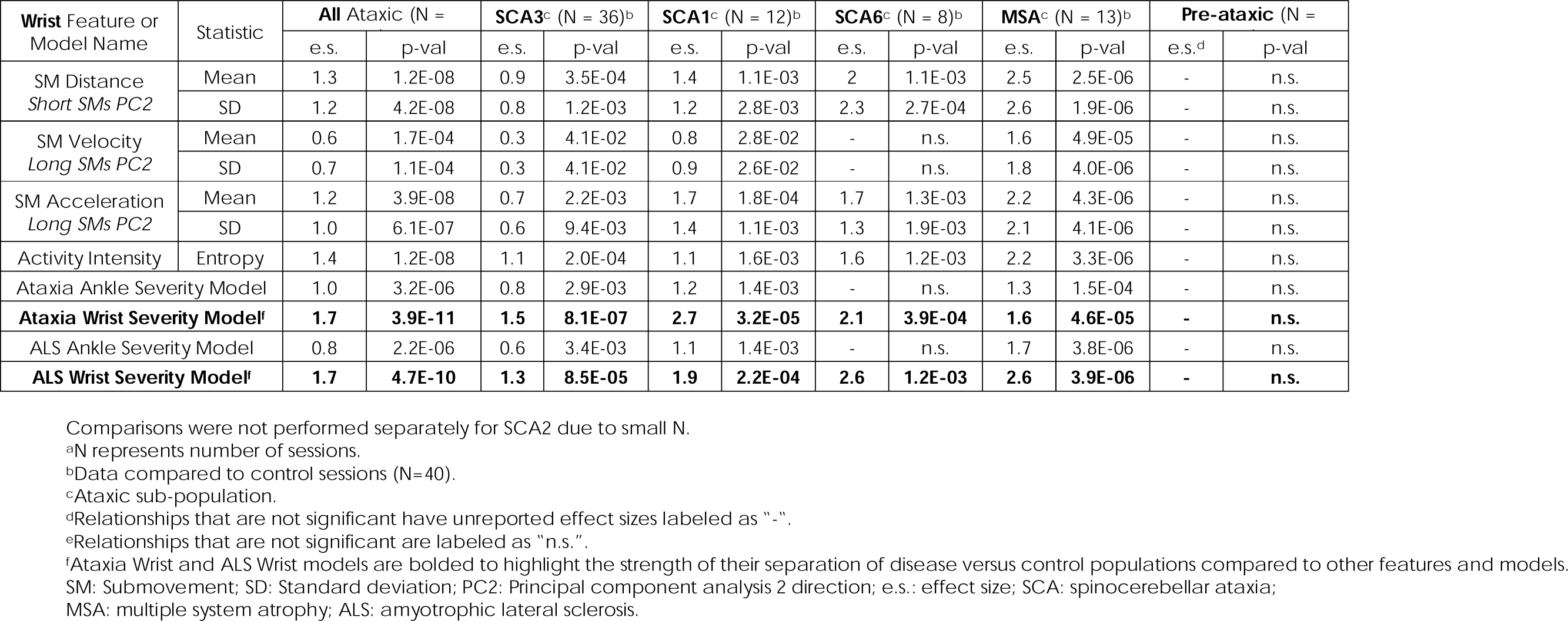
Disease versus control comparison for wrist sensor features and models.

### Cross-sectional properties of ankle movement features

Similar to the wrist, all ankle sensor-derived movement features and composite models were significantly correlated with SARA (and BARS) total score (*|r|* = 0.55 - 0.81, Table 4), SARA (and BARS) gait subscore (*|r|* = 0.52 - 0.78), PROM-Ataxia total score (*|r|* = 0.40 - 0.70), and PROM-Ataxia gait subscore (*|r|* = 0.46 - 0.72). SM distance, velocity, and acceleration all decreased with increasing motor impairment. Consistency analysis demonstrated high reliability of all ankle features and models (*ICC* = 0.90 - 0.97). As expected, the ankle-trained models, Ataxia-Ankle and ALS-Ankle, demonstrated consistently high correlations with clinical and patient-reported assessments (*|r|* = 0.65 - 0.81) and high reliability (*ICC* = 0.96 - 0.97, Table 4 and Figure 1 A-E). In addition to the ankle models, the wrist-trained ALS model (ALS-Wrist) applied to ankle data also had high correlations with clinical scales and PROMs, and demonstrated high reliability.

**Figure 1:**
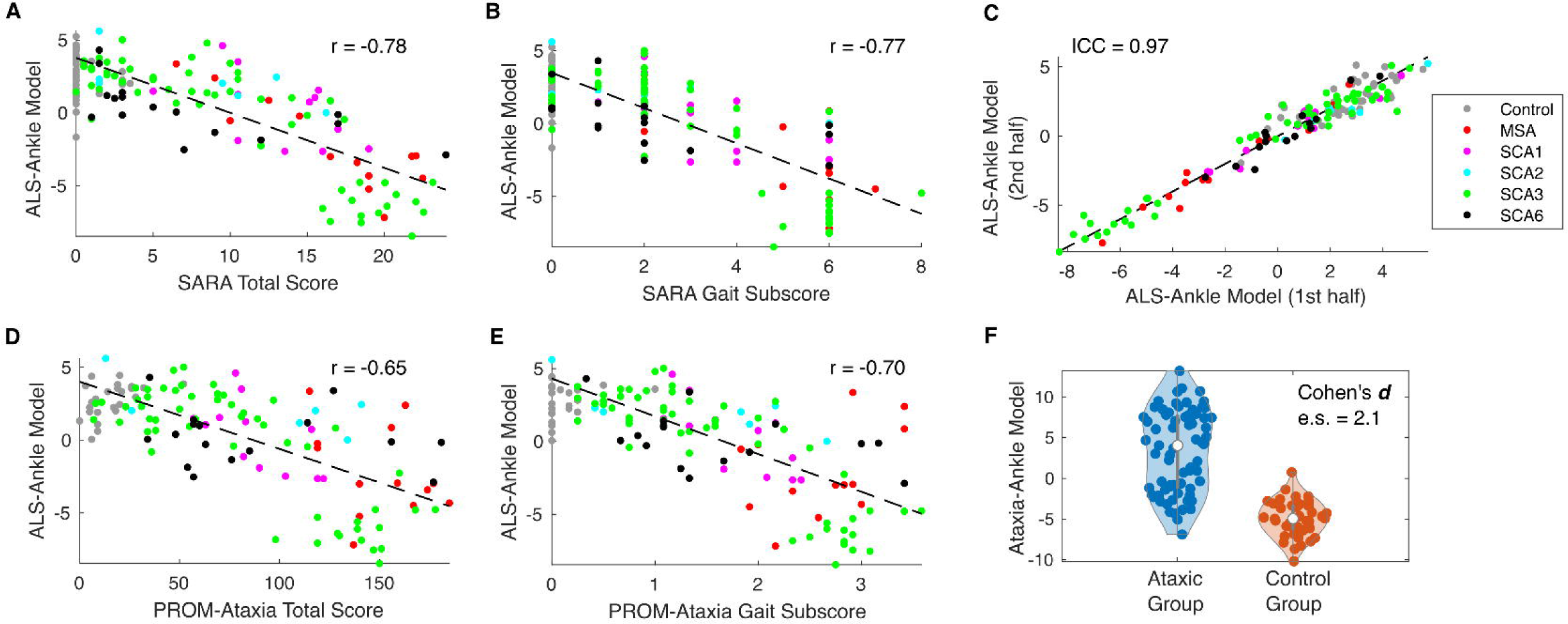
Ankle sensor measure was consistent with clinical and patient-reported assessments, reliable, and differed between ataxia and control groups. (A-B) Relationship of the ALS-Ankle model with SARA total score and gait subscore, respectively. **(C)** Test-retest reliability of the ALS-Ankle Model. **(D-E)** Relationship of the ALS-Ankle model with PROM-Ataxia total score and gait subscore, respectively. **(F)** Ataxia versus control violin plot. Each point represents a session.

**Table 4.**
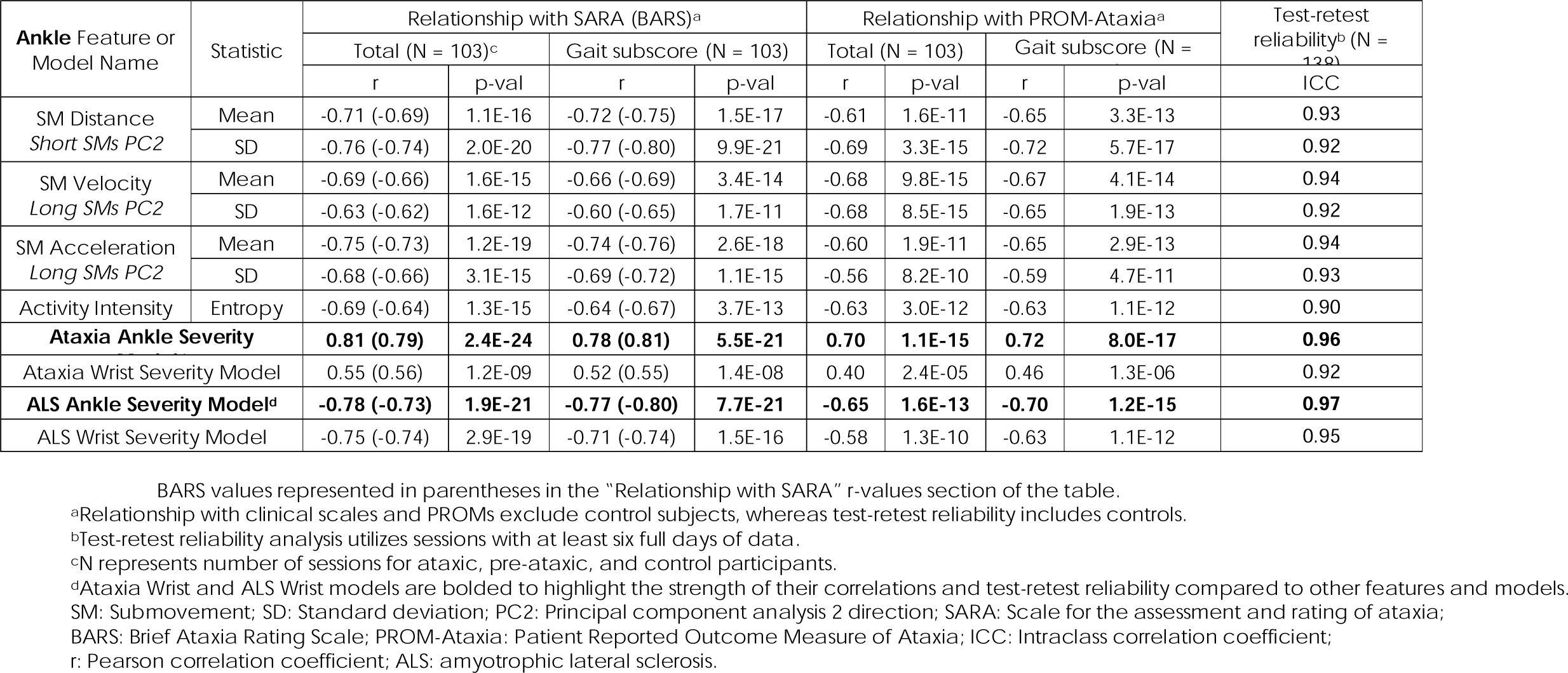
Relationship with scales and test-retest reliability of ankle sensor features and models.

Significant differences were observed between ataxic individuals and controls across all ankle features and models (*e.s.* = 1.1 - 2.1, Table 5). Group differences remained significant when considering each ataxia subpopulation separately (e.g., SCA1, SCA3, SCA6, and MSA- C). Variability in submovement acceleration was the only significantly different feature between pre-ataxic individuals and control participants after correction for multiple comparisons (Table 5). The Ataxia-Ankle model most strongly separated ataxia populations from controls (*e.s.* = 1.8 - 2.6, Table 5 and Figure 1 F), but both ALS-Ankle and ALS-Wrist models also showed strong separation (*e.s.* = 1.2 - 2.8, Table 5).

**Table 5.**
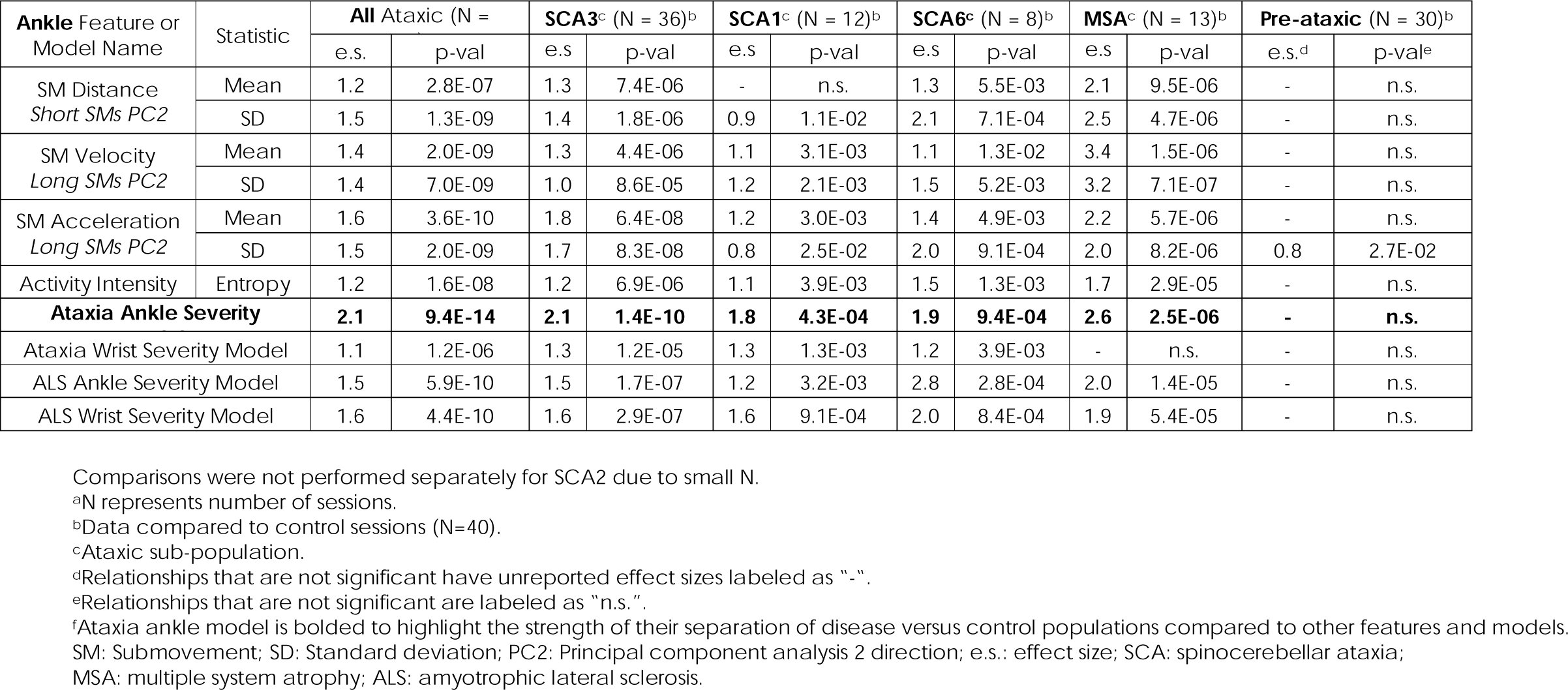
Disease versus control comparison for ankle sensor features and models.

### Longitudinal properties of wrist and ankle movement features

The majority of wrist and ankle movement features and models demonstrated statistically significant change over time (Table 6). Effect size, expressed as the mean to standard deviation ratio (|*MSDR*|) of the measure’s slopes over the population, ranged from 0.5 - 1.0. The Ataxia-Ankle and ALS-Ankle models demonstrated significant change over time when applied to ankle data, but not wrist data (Table 6). The Ataxia-Wrist and ALS-Wrist models showed significant sensitivity to disease progression when applied to ankle and wrist data, with the ALS-Wrist model having |*MSDR*| of 0.9 and 1.0 for ankle and wrist data, respectively (Table 6, Figure 2 A-D). |*MSDR*| was 0.9 for BARS total and 0.8 for SARA total (Supplementary Table 2). PROM-Ataxia total score did not show significant change, however, PROM-Ataxia gait and arm subscores were significant with |*MSDR*| of 0.8 and 0.6, respectively (Supplementary Table 2).

**Figure 2:**
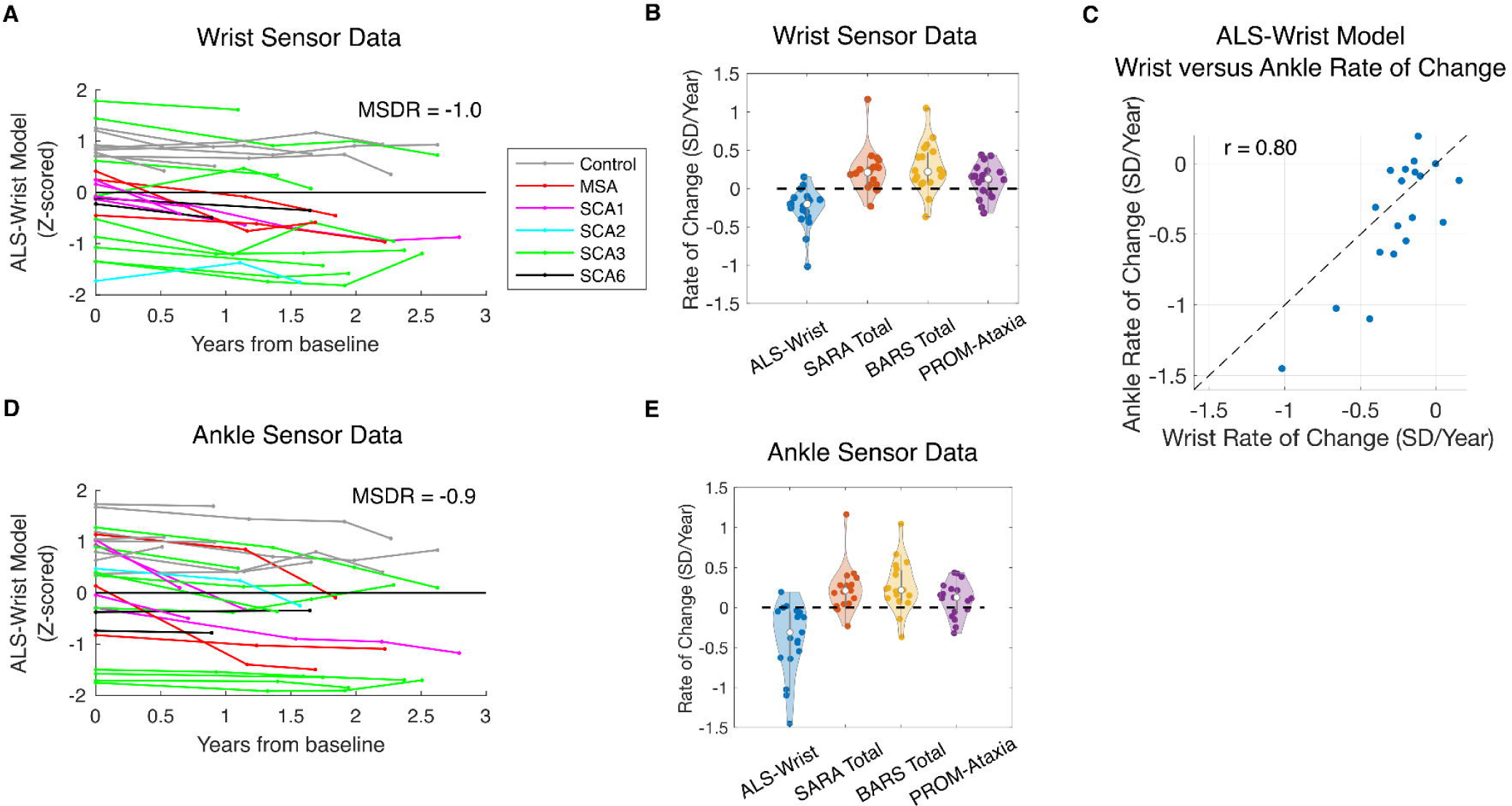
Wrist and ankle sensor measures were sensitive to disease progression. **(A)** Change over time of a wrist sensor outcome (ALS-Wrist model). **(B)** Rate of change comparison between the wrist sensor outcome (ALS-Wrist model), SARA total, BARS total, and PROM-Ataxia total. **(C)** Rate of change agreement between wrist and ankle sensor data (ALS-Wrist model applied to both). **(D)** Change over time of an ankle sensor outcome (ALS-Wrist model) **(E)** Rate of change comparison between the ankle sensor outcome (ALS-Wrist model), SARA total, BARS total, and PROM-Ataxia total. Each line or point represents an individual. For panels C-D, the ALS-Wrist model was applied to data collected from the ankle sensor.

**Table 6.** Sensitivity to disease progression for ankle and wrist sensor features and models.

| Feature or Model Name | Statistic | Ankle Sensor |  | Wrist Sensor |  | Ankle-Wrist Rate of Change |  |
| --- | --- | --- | --- | --- | --- | --- | --- |
|  |  | All Ataxic (N=19)<sup>a</sup> |  | All Ataxic |  | All Ataxic (N=19) |  |
|  |  | e.s.<sup>b,c</sup> | p-val<sup>d</sup> | e.s. | p-val | r | p-val |
| SM Distance<br>*Short SMs PC2* | Mean | -0.5 | 2.2E-02 | -0.6 | 1.4E-02 | 0.78 | 9.5E-05 |
|  | SD | -0.7 | 2.2E-02 | -0.6 | 1.6E-02 | 0.63 | 3.8E-03 |
| SM Velocity<br>*Long SMs PC2* | Mean | -0.8 | 9.4E-03 | -0.5 | 2.6E-02 | 0.70 | 8.8E-04 |
|  | SD | -0.8 | 8.3E-03 | -0.5 | 3.1E-02 | - | n.s. |
| SM Acceleration<br>*Long SMs PC2* | Mean | -0.6 | 1.6E-02 | -0.5 | 2.5E-02 | 0.76 | 1.5E-04 |
|  | SD | - | n.s. | - | n.s. | 0.64 | 3.3E-03 |
| Activity Intensity | Entropy | -0.7 | 1.6E-02 | -0.6 | 4.6E-02 | 0.56 | 1.3E-02 |
| Ataxia Ankle Severity Model |  | 0.9 | 9.1E-03 | - | n.s. | - | n.s. |
| Ataxia Wrist Severity Model |  | 0.6 | 1.1E-02 | 0.7 | 1.6E-02 | 0.79 | 6.8E-05 |
| ALS Ankle Severity Model |  | -0.6 | 1.5E-02 | - | n.s. | - | n.s. |
| **ALS Wrist Severity Model<sup>e</sup>** |  | **-0.9** | **8.0E-03** | **-1.0** | **6.0E-03** | **0.80** | **4.4E-05** |

<sup>a</sup>N represents number of ataxic participants with longitudinal data.
<sup>b</sup>Effect size is the mean progression slope divided by the standard deviation of the progression slope across participants.
<sup>c</sup>Relationships that are not significant have unreported effect sizes labeled as "-".
<sup>d</sup>Relationships that are not significant are labeled as "n.s.".
<sup>e</sup>ALS Wrist Model is bolded to highlight its strong sensitivity to disease progression compared to other features and models.
SM: Submovement; SD: Standard deviation; PC2: Principal component analysis 2 direction; e.s.: effect size;
r: Pearson correlation coefficient; ALS: amyotrophic lateral sclerosis.

Next, we evaluated whether sensor-derived movement features and composite models showed similar progression rates across the ankle and wrist sensor locations. There was significant agreement between the ankle and wrist sensors for the majority of features and models (*r* = 0.56 - 0.80, Table 6). The ALS-Wrist model, which showed the strongest progression rate effect, also showed the highest rate of change agreement between the ankle and wrist (*r* = 0.80, p < 5e-05, Figure 2 C).

## Discussion

We have shown that accelerometer data, obtained from relatively inexpensive sensors worn on the wrist and ankle during natural behavior, produce measures of motor function that strongly reflect patient-reported measures of function and clinician-performed rating scales, strongly separate ataxia from control groups, are highly reliable, and are sensitive to disease change. Wrist sensor data closely reflected both overall impairment and arm impairment, and ankle sensor data reflected overall impairment and gait impairment. Wrist and ankle movement building blocks were similarly altered, with submovements from both limbs becoming smaller and slower with disease progression. One particular composite model trained on a separate wearable sensor dataset to learn longitudinal changes in ALS (ALS-Wrist model) showed high sensitivity for capturing disease progression from both wrist and ankle data in the ataxia population.

We previously reported an interim cross-sectional analysis of this study demonstrating that accelerometer data collected from wrist and ankle sensors during natural behavior produces measures of motor function that are highly reliable and reflect patient-reported and clinician-rated motor impairment.^33^ Here, we replicate the cross-sectional analysis in a larger ataxia and control population, perform a new longitudinal analysis demonstrating the sensitivity of wearable sensor outcomes, and report promising performance of composite models trained on a separate large and longitudinal ALS dataset.^39^

Multiple studies have now shown that movement building blocks known as submovements are altered in children and adults with ataxia, both during natural behavior at home^33-34^ as well as during prescribed motor tasks such as reaching^25^ and walking.^28,38^ The submovements that compose motor behaviors become smaller and slower with increasing impairment, and are thought to reflect a hallmark characteristic of the ataxia phenotype: decomposition of movement into smaller parts.^37^ In the current study, submovement properties including distance, velocity, and acceleration were found to be smaller in ataxic individuals compared to controls. These properties were also smaller for those with greater motor impairment and became progressively smaller over time in individuals with ataxia. Mean submovement distance, velocity, and acceleration all showed significant decline over time in the longitudinal cohort. This was true for both wrist and ankle sensor data, and the rate of decline measured from participants’ wrists and ankles was strongly correlated.

We recently reported that individuals with ALS also have smaller and slower wrist and ankle submovements, derived from accelerometer data collected continuously at home.^39^ The ALS dataset consisted of a large number of individuals with ALS who had many longitudinal timepoints (188 individuals; median 15 timepoints over 1.5 years). This enabled the training of machine learning models (ALS-Wrist and ALS-Ankle) to identify patterns of change over time in ALS directly from the longitudinal sensor data, without relying on clinician scales or patient-reported data. In the ALS study, we found that overall severity estimates, produced by the wrist and ankle models, were the most sensitive for capturing disease progression in ALS.^39^ The rate of change of the sensor-based severity estimates correlated strongly with the gold standard ALS Functional Rating Scale-Revised rate of change, despite not having been trained to predict the functional rating scale. Submovement peak velocity was the most highly selected feature in the ankle and wrist ALS longitudinal models. Thus, submovement peak velocity (among other submovement properties) is reduced in both ALS and ataxias, but for different reasons. In ALS, reduced velocity may reflect muscle weakness,^52^ whereas in ataxias it may reflect movement decomposition.

The results of the current study support that patterns of submovement change are shared across ataxias and ALS. The ALS-Wrist and ALS-Ankle models demonstrated consistently high correlations with ataxia rating scales and PROM-Ataxia, had very high reliability, and strongly separated individuals with ataxia from controls. The ALS-Wrist model demonstrated the highest sensitivity to disease change for both wrist *and* ankle sensor data in the ataxia population. This was a noteworthy observation, as the ALS-Wrist model was trained on a separate dataset, in a different disease population, using a different sensor and sampling rate. This result likely reflects that there are consistencies with how ALS and ataxias progress over time at the limb submovement level, and highlights the value of the denoising effects of within-subject longitudinal modeling to remove subject-level factors and isolate changes related to disease. It also points to the generalizability of this assessment approach to other ataxias and neurological populations more broadly. While the ALS model demonstrates strong performance in ataxia, we predict that stronger sensitivity to disease progression would be obtained from a model trained on a large longitudinal dataset of individuals with ataxia, and stronger still if separate models were optimized for each genetically-defined ataxia population. In addition to model improvements, given the high reliability of the sensor outcomes, collecting data at more frequent timepoints (e.g., every 3 months) could improve sensitivity for measuring disease progression at shorter time intervals in ataxias.

The longitudinal data set was modest in size, with 19 ataxic individuals and a median of three timepoints over 1.7 years. Despite the limited timepoints to date, the high within-week reliability of the measures raised the possibility of accurately estimating the rate of change based on a few timepoints. We found that the rate of change of most wrist movement measures correlated strongly with the rate of change of the corresponding ankle feature (strongest for the ALS-Wrist model), supporting the accuracy of the rate of change estimates. We found that the ALS-Wrist model, applied to wrist and ankle sensor data, had similar sensitivity to disease change as SARA and BARS total scores. SARA and BARS scores were obtained from a single ataxia specialist; thus, inter-rater variability was not a factor in rate of change estimates for the ataxia rating scales. As expected, the effect sizes observed for SARA and BARS sensitivity in this study were substantially higher than other studies involving multiple sites.^53^ This points to an advantage of objective sensor-based outcome measures that are not susceptible to inter-rater variability and are likely to have similar properties when scaled for use in multisite interventional trials.

Low-burden wearable sensors worn at home during natural behavior are a potentially promising approach for tracking individuals with genetic ataxias in early or pre-ataxic phases of the disease. We observed one ankle sensor feature, representing variability in submovement acceleration, significantly separated pre-ataxic individuals from controls after correction for multiple comparisons. This particular measure demonstrated high reliability, but it was not one of the measures that correlated most highly with clinical scales or most strongly separated ataxic from control populations, and it did not capture disease change over time in the ataxic group. This may suggest that the informative disease-related movement patterns during natural behavior in the pre-ataxic stage may differ from the informative patterns in later stages of disease, and could reflect early adaptations or compensations for changes in motor function. The machine-learned composite models evaluated were not optimized for learning potential early patterns. Furthermore, the sensor features selected a priori for analysis were based largely on prior data from ataxic individuals. Future work in a larger cohort of pre-ataxic individuals is needed to further understand early movement changes and their evolution over time, as well as to train models optimized for this task.

## Limitations

There are some limitations to this study. The rare disease cohort studied included a relatively large cross-sectional population, with a smaller set of individuals having longitudinal data. This limited the ability to independently evaluate longitudinal properties within genetically defined cohorts, as well as the pre-ataxic group. More longitudinal time points are also needed in each individual to produce reliable estimates of ataxia rating scale and PROM rate of change, which would then enable an assessment of rate of change correlations between clinical assessments and sensor-based assessments. Expanded longitudinal data would also support a detailed understanding of factors underlying differences in wrist and ankle sensor rate of change and how best to combine data from both limbs to produce an optimized overall severity estimate. These limitations will be addressed with ongoing longitudinal data collection in this cohort. Additionally, a single clinician rater evaluated all participants, which may have resulted in an overestimation of the ataxia rating scale’s sensitivity to change in a clinical trial setting.

## Data availability

Data included in this study will be shared upon request from qualified investigators.

## Data Availability

Data included in this study will be shared upon request from qualified investigators.

## Acknowledgments

The authors thank Anna Luddy for assistance with recruitment, and Jessey Ouillon and Jeremy Edgerton for contributions on the original cross-sectional cohort. The authors would also like to thank the participants and families who were involved in this project.

## Funding

This work was supported by the National Institutes of Health (R01 NS117826 and R01 NS134597) and the Friedreich’s Ataxia Research Alliance.

## Competing interests

For the methods for extracting and characterizing submovements from wearable sensor data, a PCT (US2022/081374) was filed on December 12, 2022, titled “System and method for clinical disorder assessment”. An earlier US Provisional Application (Serial No. 63/288,619) was filed on December 12, 2021. The patent applicant is the institution (Massachusetts General Hospital) and inventor is Anoopum Gupta. The remaining authors declare no competing interests.

## Supplementary material

Supplementary material is available at *Brain* online.

## Notes

### Funding Statement

This work was supported by the National Institutes of Health (R01 NS117826 and R01 NS134597) and the Friedreichs Ataxia Research Alliance.

### Author Declarations

The study was approved by the Partners Healthcare Research Committee Institutional Review Board (no. 2019P003458). Age-matched control data collected under a separate study approved by the same review board above (no. 2019P002752) was also used in this analysis to increase the size of the control cohort.

