## Supplementary Tables and Methods for "At-home wearables and machine learning capture motor impairment and progression in adult ataxias"

**Supplementary Information**

**Supplementary Tables and Figures**

**Supplementary Table 1. Participant demographic and clinical information.** FA study control participants did not complete BARS, SARA, or PROM-Ataxia. All participants' BARS, SARA, and PROM-Ataxia scores were from their initial session. The walking devices used most frequently across a participant’s longitudinal time points were reported. Their most recent session’s walking device was used if only two sessions were completed. BARS, Brief Ataxia Rating Scale; SARA, Scale for the Assessment and Rating of Ataxia; PROM-Ataxia, Patient-Reported Outcome Measure of Ataxia; *n*: number.

| **Age Group** | **Subject** | **Sex** | **Diagnosis** | ***n* Timepoints** | **Span (Years)** | **BARS Total** | **SARA Total** | **PROM-Ataxia Total** | **Walking Device** |
| --- | --- | --- | --- | --- | --- | --- | --- | --- | --- |
| 28-40 | 1 | F | SCA3 | 4 | 2.4 | 14.5 | 16 | 130 | Walker |
| 28-40 | 2 | F | Pre-A (SCA2) | 1 | 0 | 4 | 1.5 | 13 | None |
| 28-40 | 3 | F | SCA1 | 2 | 0.6 | 6 | 9.5 | 78 | None |
| 28-40 | 4 | F | SCA1 | 1 | 0 | 7 | 5 | 57 | None |
| 28-40 | 5 | M | Pre-A (SCA6) | 1 | 0 | 1 | 1.5 | 127 | Cane |
| 28-40 | 6 | M | Pre-A (SCA6) | 1 | 0 | 1.5 | 2 | 114 | Cane |
| 28-40 | 7 | M | Control | 1 | 0 | N/A | N/A | N/A | None |
| 28-40 | 8 | F | SCA3 | 1 | 0 | 21 | 22.5 | 98 | Rollator |
| 28-40 | 9 | M | Control | 1 | 0 | N/A | N/A | N/A | None |
| 28-40 | 10 | M | Control | 1 | 0 | N/A | N/A | N/A | None |
| 28-40 | 11 | F | Control | 3 | 1.7 | 0 | 0 | 19 | None |
| 28-40 | 12 | M | SCA3 | 3 | 1.7 | 6 | 3.5 | 56 | None |
| 28-40 | 13 | M | Control | 1 | 0 | N/A | N/A | N/A | None |
| 28-40 | 14 | F | Pre-A (SCA3) | 3 | 1.8 | 0.5 | 1 | 72 | None |
| 28-40 | 15 | F | Pre-A (SCA3) | 1 | 0 | 0 | 1 | 34 | None |
| 28-40 | 16 | F | Pre-A (SCA3) | 4 | 2.6 | 0 | 0 | 58 | None |
| 28-40 | 17 | F | Control | 1 | 0 | N/A | N/A | N/A | None |
| 41-50 | 18 | F | SCA2 | 3 | 1.6 | 9 | 10.5 | 110 | None |
| 41-50 | 19 | M | SCA3 | 1 | 0 | 5.5 | 6 | 81 | None |
| 41-50 | 20 | F | Control | 1 | 0 | N/A | N/A | N/A | None |
| 41-50 | 21 | M | SCA6 | 2 | 0.9 | 8 | 7 | 57 | Walking stick |
| 41-50 | 22 | F | Control | 2 | 0.9 | 0 | 0 | 6 | None |
| 41-50 | 23 | M | SCA3 | 3 | 1.9 | 17 | 18.5 | 148 | Wheelchair |
| 41-50 | 24 | M | Control | 1 | 0 | N/A | N/A | N/A | None |
| 41-50 | 25 | M | Control | 1 | 0 | N/A | N/A | N/A | None |
| 41-50 | 26 | M | SCA3 | 1 | 0 | 18.5 | 22 | 150 | Motorized chair |
| 41-50 | 27 | M | Control | 2 | 0.5 | N/A | N/A | N/A | None |
| 41-50 | 28 | F | Pre-A (SCA3) | 4 | 2.1 | 3.5 | 3 | 52 | None |
| 41-50 | 29 | M | Control | 1 | 0 | N/A | N/A | N/A | None |
| 41-50 | 30 | F | SCA3 | 4 | 2.5 | 15 | 17 | 119 | Walker |
| 41-50 | 31 | F | Control | 2 | 0.5 | N/A | N/A | N/A | None |
| 41-50 | 32 | M | Control | 1 | 0 | N/A | N/A | N/A | None |
| 41-50 | 33 | F | SCA1 | 1 | 0 | 13.5 | 15 | 116 | Rollator |
| 41-50 | 34 | F | Control | 4 | 2.2 | 0 | 0 | 9 | None |
| 41-50 | 35 | M | Pre-A (SCA3) | 4 | 2.3 | 4 | 5 | 23 | None |
| 41-50 | 36 | F | Control | 1 | 0 | 0 | 0 | 14 | None |
| 51-60 | 37 | F | SCA3 | 4 | 2.6 | 7.5 | 10.5 | 69 | Cane |
| 51-60 | 38 | M | SCA1 | 2 | 1.1 | 15.5 | 15.5 | 70 | Walker |
| 51-60 | 39 | F | MSA | 3 | 2.2 | 19 | 18 | 174 | Rollator |
| 51-60 | 40 | F | Control | 1 | 0 | 0 | 0 | 6 | None |
| 51-60 | 41 | M | SCA3 | 2 | 1.1 | 12 | 10.5 | 31 | None |
| 51-60 | 42 | M | SCA2 | 1 | 0 | 16.5 | 16 | 134 | Rollator |
| 51-60 | 43 | F | Control | 1 | 0 | 1 | 1.5 | 0 | None |
| 51-60 | 44 | M | Pre-A (SCA3) | 3 | 1.6 | 1 | 0.5 | 42 | None |
| 51-60 | 45 | F | SCA3 | 2 | 1.4 | 8.5 | 12 | 55 | Walker |
| 51-60 | 46 | F | SCA3 | 1 | 0 | 6 | 7.5 | 67 | None |
| 51-60 | 47 | F | SCA6 | 2 | 1.6 | 5.5 | 5 | 48 | None |
| 51-60 | 48 | M | SCA3 | 4 | 2.3 | 12.5 | 9 | 20 | None |
| 51-60 | 49 | M | MSA | 1 | 0 | 16 | 19 | 185 | Gait belt |
| 51-60 | 50 | F | SCA3 | 1 | 0 | 13.5 | 17.5 | 114 | Motorized chair |
| 51-60 | 51 | M | Control | 4 | 2.6 | 0 | 0 | 32 | None |
| 61-70 | 52 | F | SCA6 | 1 | 0 | 21 | 24 | 177 | Wheelchair |
| 61-70 | 53 | F | Pre-A (SCA6) | 1 | 0 | 1 | 1.5 | 35 | None |
| 61-70 | 54 | M | SCA6 | 1 | 0 | 14.5 | 12 | 54 | Walking stick |
| 61-70 | 55 | M | Pre-A (SCA6) | 2 | 1.1 | 2.5 | 1 | 63 | None |
| 61-70 | 56 | F | SCA6 | 1 | 0 | 14.5 | 17 | 156 | Walker |
| 61-70 | 57 | M | SCA3 | 1 | 0 | 12 | 12 | 160 | Rollator |
| 61-70 | 58 | M | MSA | 1 | 0 | 18.5 | 22 | 159 | Walker |
| 61-70 | 59 | F | Control | 1 | 0 | 0 | 0 | 52 | None |
| 61-70 | 60 | F | SCA1 | 4 | 2.8 | 10.5 | 10.5 | 90 | Walker |
| 61-70 | 61 | F | Control | 1 | 0 | 0 | 0 | 28 | None |
| 61-70 | 62 | F | MSA | 3 | 1.8 | 7 | 6.5 | 115 | Walker |
| 61-70 | 63 | F | MSA | 1 | 0 | 14.5 | 16.5 | 140 | Wheelchair |
| 61-70 | 64 | M | MSA | 3 | 1.7 | 15 | 14.5 | 119 | Walker |
| 61-70 | 65 | M | Control | 4 | 2.3 | 1.5 | 3.5 | 18 | None |
| 61-70 | 66 | M | SCA3 | 2 | 1.7 | 17.5 | 18 | 150 | Walker |
| 61-70 | 67 | M | Pre-A (SCA6) | 3 | 1.7 | 6 | 3 | 57 | None |
| 61-70 | 68 | M | MSA | 1 | 0 | 8.5 | 10 | 119 | None |
| 61-70 | 69 | F | SCA3 | 1 | 0 | 3.5 | 3 | 50 | None |
| 61-70 | 70 | M | Control | 1 | 0 | N/A | N/A | N/A | None |
| 71-80 | 71 | F | SCA1 | 2 | 0.7 | 14 | 10.5 | 83 | Walker |
| 71-80 | 72 | F | SCA6 | 1 | 0 | 17 | 17 | 85 | Rollator |
| 71-80 | 73 | M | Control | 1 | 0 | N/A | N/A | N/A | None |
| 71-80 | 74 | F | Pre-A (SCA2) | 2 | 1.3 | 1 | 1.5 | 31 | None |
| 71-80 | 75 | F | Control | 2 | 0.9 | 0.5 | 0.5 | 26 | None |
| 71-80 | 76 | F | SCA3 | 1 | 0 | 16 | 16.5 | 128 | None |

**Supplementary Table 2.** Sensitivity to disease progression for

clinical scales and patient-reported outcome measures (PROMs).^a^N represents number of ataxic participants with longitudinal data.

^b^Effect size is the mean progression slope divided by the standard deviation of progression slope.

^c^Relationships that are not significant are labeled as “n.s.”.

SARA: Scale for the assessment and rating of ataxia; BARS: Brief Ataxia Rating Scale;

PROM-Ataxia: Patient Reported Outcome Measure of Ataxia; e.s.: effect size.

| **Clinical Scale/PROM** | **All Ataxic** (N=19)^a^ | |
| --- | --- | --- |
|  | e.s.^b^ | p-val |
| SARA Total | 0.8 | 1.0E-02 |
| BARS Total | 0.9 | 7.6E-03 |
| PROM-Ataxia Total | - | n.s.^c^ |
| SARA Gait Subscore | - | n.s. |
| BARS Gait Subscore | 0.4 | 5.0E-02 |
| PROM-Ataxia Gait Subscore | 0.8 | 6.7E-03 |
| SARA Arm Subscore | 0.7 | 2.8E-02 |
| BARS Arm Subscore | 0.8 | 3.5E-02 |
| PROM-Ataxia Arm Subscore | 0.6 | 3.3E-02 |

**Supplementary Methods**

**Recruitment and Consent**

All but one individual with SCA had genetic confirmation of their condition: one participant had a first-degree relative with genetic confirmation and a phenotype consistent with their diagnosis. All but one MSA-C participant had a probable diagnosis: one participant had a possible diagnosis of MSA-C or atypical parkinsonism, but the participant’s neurologist thought MSA-C was more likely.

**Virtual Appointment and Neurological Assessment**

Four missing scores were SARA heel-to-shin (left and right): two were unable to be scored due to poor task performance, while the other two couldn’t be scored due to technical difficulties (poor video quality, etc). One missing score was SARA finger-to-nose right, which was not performed for an unknown reason. Nine missing scores were SARA finger-chase right: seven were unable to score due to poor task performance, one was not performed due to environmental constraints, and one was not performed for an unknown reason. Eight missing scores were from SARA finger-chase left: seven were unable to be scored due to poor performance, and one was not performed due to environmental constraints. Two missing scores were from SARA alternating hand movements (one right and one left): both were unable to be scored due to technical difficulties.

Eight BARS scores for individual tasks were missing across all participants and timepoints. Two missing scores were from BARS gait, with both not performed due to safety concerns. Four missing scores were from BARS heel-to-shin (right and left), with two unable to be scored due to poor performance and the other two unable to be scored due to technical difficulties. One missing score was from BARS finger-nose right and was not performed for an unknown reason. One missing score was from BARS oculomotor and couldn’t be scored due to technical difficulties.
